# A single composite index of semantic behavior tracks symptoms of psychosis over time

**DOI:** 10.1101/2025.03.05.25323232

**Authors:** Claudio Palominos, Maryia Kirdun, Amir H. Nikzad, Michael Spilka, Philipp Homan, Iris E. Sommer, Sunny X. Tang, Wolfram Hinzen

## Abstract

Semantic variables automatically extracted from spontaneous speech characterize anomalous semantic associations generated by groups with schizophrenia spectrum disorders (SSD). However, with the use of different language models and numerous aspects of semantic associations that could be tracked, the semantic space has become very high-dimensional, challenging both theoretical understanding and practical applications. This study aimed to summarize this space into a single composite semantic index and to test whether it can track diagnosis and symptom profiles over time at an individual level. The index was derived from a principal component analysis (PCA) yielding a linear combination of 117 semantic variables. It was tested in discourse samples of English speakers performing a picture description task, involving a total of 103 individuals with SSD and 36 healthy controls (HC) compared across four time points. Results showed that the index distinguished between SSD and HC groups, identified transitions from acute psychosis to remission and stabilization, predicted the sum of scores of the Thought, Language and Communication (TLC) index as well as subscores, capturing 65% of the variance in the sum of TLC scores. These findings show that a single indicator meaningfully summarizes a shift in semantic associations in psychosis and tracks symptoms over time, while also pointing to variance unexplained, which is likely covered by other semantic and non-semantic factors.

## 1. Introduction

Disorganized speech is a hallmark of psychotic discourse and remains one of the key clinical criteria for diagnosing psychosis (Palaniyappan, 2022). It manifests in derailment, incoherence and loss of goal, contributing to the broader positive symptoms characteristic of psychotic disorders. Often linked to disruptions in semantic processing – the ability to understand and produce meaning in language – these impairments play a critical role in coherent communication. Semantic associations are especially important for building and maintaining logical connections in discourse, a notion that has sparked considerable clinical and research interest since Bleuler’s initial formulation of schizophrenia (1911). Subsequent research, such as Andreasen (1979, 1986), attempted to manualize assessment of disorganized speech through tools like the Thought, Language, and Communication (TLC) scale.

More recently, natural language processing (NLP) has offered tools to automatically extract semantic features based on large language models (LLMs), with promising performance in assessing disrupted semantic associations (Tang et al, 2021; Voppel et al, 2021; Figueroa-Barra et al., 2022; Alonso-Sánchez et al., 2023; Arslan et al., 2024). Semantic variables are often calculated based on the distributional semantics hypothesis (Harris, 1954; Firth, 1957; Miller & Charles, 1991), which suggests higher semantic similarity between words that are co-occurring in similar contexts. Distributional properties of linguistic units (i.e., words or sub-words) in large linguistic corpora are then represented computationally as dense, high-dimensional vectors known as word embeddings (Mikolov et al., 2013a; Pennington et al., 2014; Grave et al., 2018; Devlin et al., 2019). Such representations allow calculation of semantic similarity through mathematical operations such as cosine distance between two given vectors.

Today, various language models have been used in the context of speech in psychosis, each with different model-architecture and training corpora. Among them are shallow neural network models like word2vec (Mikolov et al., 2013b), which treat words as atomic units, and fastText (Grave et al., 2018), which represents words as bags of n-grams or subword units. More advanced models include BERT (Devlin et al., 2019), a transformer-based model trained on bidirectional contextual embeddings, and the GPT family, which generates text autoregressively using transformer architectures. Since semantic similarity features are extractible from any given model with multiple approaches, there has been a significant increase in similarity metrics used for classification of patients with psychosis from controls (Bedi et al., 2015; Voppel et al., 2021; Çabuk et al., 2024; Arslan et al., 2024). Model-dependency of semantic similarity measures and multi-algorithmic strategies to compute them, hinder consistent interpretation of results within what has become a very high-dimensional variable space.

In addition, different semantic similarity metrics relate differently to clinical or cognitive correlates (Tang et al., 2022; He et al., 2024a; He et al., 2024b; Alonso-Sánchez et al., 2023; Palominos et al., 2024), often creating inconsistencies in the way that similarity metrics relate to clinical language disturbances. For example, early studies since Elvevåg et al. (2007) have suggested that greater incoherence in speech would translate into lower values in mean semantic similarity (see also Morgan et al., 2022). But more recently, a pattern of *increased* mean semantic similarity in psychosis has been documented across different languages, disease stages, and tasks (Pintos et al., 2022; Alonso-Sánchez et al., 2023; Parola et al., 2023; He et al., 2024a; Çabuk et al., 2024; Arslan et al., 2024). This phenomenon has been conceptualized as a ‘shrinking’ of the semantic space, where the scope of semantic differences becomes more constrained (He et al., 2024a; Palominos et al., 2024). An emergent subfield has related such semantic and other probabilistic LLM-based measures to both brain structure and function, sometimes demonstrating their greater sensitivity over traditional clinical or neuropsychological metrics in this respect (Palaniyappan et al., 2019; Liang et al., 2022; Limongi et al., 2023; Sharpe et al., 2024; Alonso-Sánchez et al., 2024). These findings reinforce the potential of LLM-based language features for advancing patient-centered psychiatric assessments and for their neurobiological grounding.

As things stand in this field, a clear theoretical foundation for why certain semantic variables offer more explanatory power than others, and under which conditions, remains missing. For example, it remains unclear why second-order semantic similarities (between two words as separated by another) might be more sensitive than the first order ones (between consecutive words) (Parola et al., 2023), or how dynamic and static variables might contribute differently to our understanding of semantic associations (Palominos et al., 2024). This highlights the need for a reliable approach reducing the high-dimensional semantic space to a lower-dimensional one, in a way that would retain interpretability and be of clinical utility.

Here we reasoned that semantic associations are characterized by a large range of computational semantic variables, all of which may contribute to understanding the variance involved. To characterize this variance in patients and controls, we constructed an index derived from a principal component analysis (PCA) of features based on the semantic similarity concept. This index is conceived as an agnostic tool, which captures the semantic patterns that each variable reveals in relation to psychosis and its symptoms. PCA is one of the oldest multivariate statistical techniques, which allows to significantly reduce the dimensionality of data (Jolliffe, 2002). It explores the relationships between features and transforms correlated variables into a set of uncorrelated variables using a covariance matrix (Nardo et al., 2005). The goal of PCA is to compress the original size of data while capturing the maximum variances in the features, irrespective of their relevance to external factors, such as clinical context. New variables, called principal components, are computed as linear combinations of the original variables, with the first principal component having the largest possible variance (Abdi & Williams, 2010).

The aim of this study was to test this PCA-based index as a suitable summary indicator of the semantic behavior of an individual in its multiple dimensions, and to assess its explanatory power and ability to track both group differences and changing symptoms over time. Our three specific objectives were: (i) to determine whether a composite index of cross-model semantic similarity metrics can distinguish between psychotic patients and healthy controls; (ii) to understand longitudinal changes in the calculated composite index over time, particularly as patients transition from an acute psychotic stage to remission; and (iii) to assess whether this index can predict various symptoms as measured by clinical symptom scales. Achieving these objectives would mean that a semantic index automatically calculated for each individual would provide an independent and single score reflecting linguistic behavior at a semantic level along with psychotic symptoms.

## 2. Materials and methods

### 2.1 Participants

Individuals with schizophrenia spectrum disorders and healthy volunteers were recruited from inpatient and outpatient facilities and two different cohorts (LPoP and Remora) at the Zucker Hillside Hospital in Glen Oaks, New York. The samples together covered up to four time points in total, at which speech was collected. For the purpose of this analysis, seven SSD and two HC participants were excluded either due to recording issues or missing transcripts for the first timepoint. The first session, referred to here as T1 (timepoint 1), took place at an acute psychiatric unit, as soon as possible after the participant was deemed sufficiently stable to participate in research, in the case of the LPoP cohort, and either during admission or as a stable outpatient, in the case of Remora. The second session (T2) for LPoP participants was held after one to three weeks, at stabilization or discharge from the hospital. The range of 1-3 weeks was set to limit variability in follow-up time. If participants were discharged prior to 1 week, then the second timepoint occurred soon after discharge, and if the admission lasted longer than 3 weeks (often due to housing and dispositional issues), the participant was assessed in advance of their discharge at the third week mark. The third follow-up assessment was 3 months (T3) after the second timepoint, and the fourth one after 6 months (T4). The final analysis included speech samples from 139 participants (SSD = 103 and HC = 36) for T1; 47 SSD participants for T2; 57 participants (SSD = 33; HC = 24) for T3 and 13 SSD participants for T4.

Table 1 shows all the time points as well as how the participants from the two cohorts were grouped for comparability in our analysis, based on when their assessments took place. LPoP participants were between 18 and 40 years old on admission, with a current psychotic episode or significant positive or disorganized symptoms of psychosis. Remora healthy volunteers were between 21 and 38 years old, while the patients were between 18 and 51 years old and they were both inpatients and outpatients, which means they were more mixed compared to LPoP.

**Table 1:**
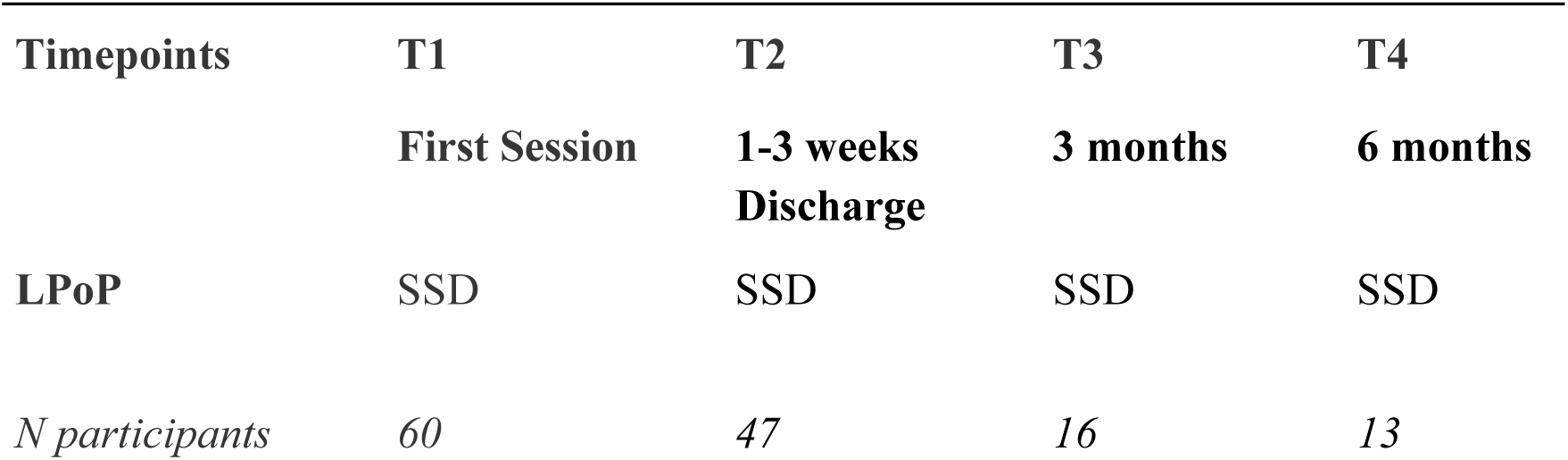
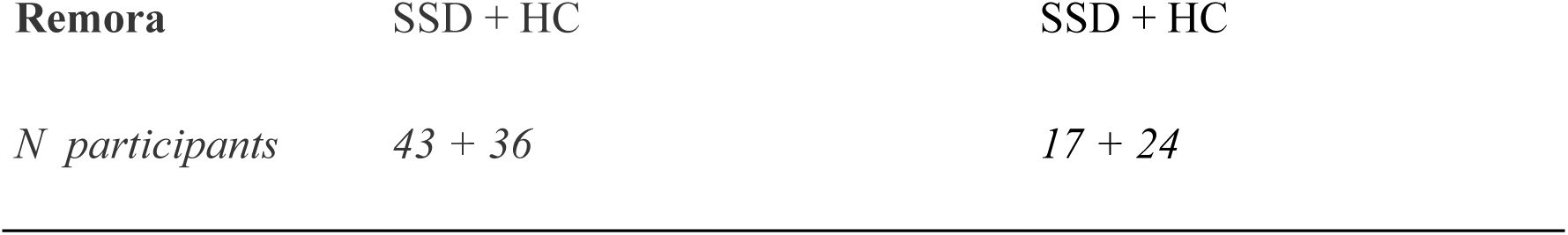
Participants from LPoP and Remora across 4 sessions.

Samples from the two cohorts were formed for each of two main analyses separately, as specified in Tables 2 and 3, respectively. The initial index calculation included all available SSD participants, in order to maximize the number of observations and variance. In the first main analysis, group comparisons were then performed between LPoP SSD and Remora HC participants (Table 2), to capture group and time effects. As our focus was on patients transitioning from an acute to a remitted stage, Remora SSD participants were excluded from this analysis due to the variability of acuity at the first timepoint, while LPoP SSD participants were all in an acute stage. To assess the evolution of the index over time and its relationship with symptoms, the LPoP-only SSD cohort was analyzed (Table 3). There were no significant differences in demographic characteristics between the time points in the case of LPoP-only, however there were differences in TLC scores. Age, education and sex differences were controlled for in the statistical analyses by including them as covariates in all mixed-effects linear models.

**Table 2:** Demographic Characteristics LPoP SSD + Remora HC.

|  | **T1** |  |  | **T3 (3 months)** |  |  |
| --- | --- | --- | --- | --- | --- | --- |
|  | **SSD** | **HC** | **p-value** | **SSD** | **HC** | **p-value** |
| *n* | 60 | 36 |  | 16 | 24 |  |
| **Age** | 25.90 (7.65) | 29.15 (7.1) | <0.001*** | 27.50 (8.08) | 32.50 (8.03) | 0.08 |
| **Sex** |  |  | — |  |  | — |
| Female | 19 | 19 |  | 1 | 15 |  |
| Male | 40 | 17 |  | 15 | 9 |  |
| Intersex | 1 |  |  |  |  |  |
| **Education** | 15.00 (3.00) | 16.00 (2.00) | <0.001*** | 15.00 (0.25) | 17.00 (2.00) | <0.001*** |
| **TLC total** | 18.00 (23.25) | 2.00 (3.25) | <0.001*** | 7.00 (10.75) | 2.00 (4.00) | <0.01** |
| **SANS total** | 27.00 (13.25) | 2.00 (5.50) | <0.001*** | 21.00 (17.75) | 4.00 (6.50) | <0.001** |
| **BPRS total** | 49.00 (13.50) | 21.00 (3.50) | <0.001*** | 43.00 (21.50) | 20.00 (5.50) | <0.001*** |

The significant associations are tagged by asterisks (\*<0.05, \*\*<0.01, and \*\*\*<0.001). Age, education and TLC, SANS, BPRS total scores are reported by medians and interquartile ranges (IQRs). P-values were calculated using the Mann-Whitney U test. Sex was excluded from calculation due to low counts for ‘Intersex’ and ‘Female’ categories. TLC - Scale for the Assessment of Thought Language and Communication. SANS - Scale for the Assessment of Negative Symptoms. BPRS - Brief Psychiatric Rating Scale.

**Table 3:** Demographic Characteristics LPoP.

|  | **T1** | **T2** | **T3** | **T4** | **p-value** |
| --- | --- | --- | --- | --- | --- |
| *n* | 60 | 47 | 16 | 13 |  |
| **Age** | 25.90 (7.65) | 25.30 (6.65) | 27.50 (8.08) | 25.70 (5.30) | 0.7 |
| **Sex** |  |  |  |  | — |
| Female | 19 | 14 | 1 | 2 |  |
| Male | 40 | 32 | 15 | 11 |  |
| Intersex | 1 | 1 |  |  |  |
| **Education** | 15.00 (3.00) | 15.00 (3.00) | 15.00 (0.25) | 15.00 (1.00) | 0.9 |
| **TLC total** | 18.00 (23.25) | 14.00 (15.50) | 7.00 (10.75) | 10.00 (12.00) | <0.01** |
| **SANS total** | 27.00 (13.25) | 24.50 (13.75) | 21.00 (17.75) | 22.00 (28.00) | 0.3 |
| **BPRS total** | 49.00 (13.50) | 40.50 (19.00) | 43.00 (21.50) | 31.00 (10.00) | <0.001*** |

Age, education, SANS, TLC, and BPRS total scores are reported by median (IQR). P-values were calculated using the Kruskal-Wallis test. Sex was excluded from calculation due to low counts for ‘Intersex’ and ‘Female’ categories.

### 2.2 Materials and procedure

Participants in both studies were recorded while providing their responses to different tasks (fluency and journaling tasks, picture descriptions and paragraph reading). For the present study, speech samples from the first picture description task were extracted to be assessed. The picture description task was chosen as this task provides naturalistic speech, which is nonetheless constrained (or anchored) in an external stimulus, thereby restricting variance, unlike free speech. To ensure consistency across the LPoP and Remora cohorts, similar pictures were used. These pictures depicted common, everyday scenes (e.g., people engaged in various activities in a living room, kitchen, market, celebrating a birthday). The choice of pictures aimed to allow for a variety of responses while maintaining a similar stimulus across participants. All healthy volunteers were assessed virtually through a video call software. Recordings were transcribed by human annotators, de-identified, and reviewed for accuracy. All transcripts featured utterance boundaries that were determined based on pauses and syntactic completeness.

To assess psychosis symptoms, participants underwent clinical semi-structured interviews for the DSM-IV (First & Gibbon, 2004) and met DSM-5 criteria for SSD. The Scale for the Assessment of Thought Language and Communication (TLC) (Andreasen, 1986) and the Scale for the Assessment of Negative Symptoms (SANS) were used for clinical language ratings. Healthy volunteers were confirmed to be free of current psychiatric disorders. Among exclusion criteria were substance-induced disorders, intellectual disability (IQ<70), autism spectrum disorder, language or speech disorders, physical impairments affecting speech, serious neurological or endocrine disorders, and any condition or treatment affecting brain or language function, as well as any factor preventing informed consent. Participants were evaluated with the WRAT (Wide Range Achievement Test, reading test) to assess reading proficiency in English. All participants signed a written informed consent. The research procedures were approved by the Institutional Review Boards at the Feinstein Institutes for Medical Research, Northwell Health.

### 2.3 LLM embeddings

Three pre-trained language models were used to generate similarity measures. For word-level analysis, we used the static fastText (Grave et al., 2018) model and contextual BERT (Bidirectional Encoder Representations from Transformers, Devlin et al., 2019), utilizing the pre-trained model for English ‘bert-base-cased’, fine-tuned for each sample; while at sentence-level, a SentenceTransformers model was employed (Reimers & Gurevych, 2019), using specifically the ‘all-mpnet-base-v2’ model. Compared to other static models, fastText provides more flexibility, covering out-of-vocabulary words, considering subword information and incorporating morphological information into word representation (Bojanowski et al., 2017). Although its static nature may limit the model’s awareness of context, fastText shows effectiveness in the studies of semantics in psychosis (Pintos et al., 2022; Choe et al., 2023; He et al., 2024, Palominos et al., 2024). Opposite to fastText, BERT (trained on the English Wikipedia and the GoogleBooks Corpus) provides more precise and nuanced word representations that change depending on the context surrounding the word. Instead of processing text sequentially, BERT approaches the context of words in an utterance bi-directionally. The combination of static (fastText), contextualized (BERT) word embeddings and sentence-level representations (SentenceTransformers) helped to ensure a comprehensive analysis that captures both word-level and sentence-level semantics. The selected models have demonstrated strong performance in prior research and semantic analysis in psychosis (He et al., 2024a; Alonso-Sánchez et al., 2024; Palominos et al., 2024), making them reliable choices for this task. Moreover, incorporating models with distinct architectures and training methodologies allowed for a more diverse set of embeddings, minimizing biases that might arise from relying on a single type of model.

### 2.4 Data analysis and semantic similarity measures

Speech transcripts were pre-processed and cleaned in Python so as to remove any irrelevant information (about the interviewer, speaker, clusters, etc.) and extra symbols or whitespaces. The texts were normalized and tokenized with spaCy (version 3.7.4), using the ‘en_core_web_sm’ model for English. The list of default stop words in spaCy was updated accordingly for the task. The stop word lists can be found in the supplementary materials (SM). The three language models mentioned above were used to calculate embeddings on the word and sentence levels for each participant. Only content words were embedded in fastText. In order to retain contextual and syntactic nuances in word representations, BERT embeddings were first calculated using complete sentences that included punctuation and stop words. Special tokens and punctuations were then dropped for semantic similarity calculation. Sentence embeddings were calculated using SentenceTransformers for the sentences within the boundaries previously determined by the annotators. Once all the embeddings were generated, cosine semantic similarities between content words, between all words and between sentences were calculated. Finally, based on semantic similarities, a total of 117 dynamic and static semantic features were computed in order to perform the PCA analysis. Feature selection was motivated by the desire to include a comprehensive list of variables covering the semantic variable space as used in previous computational semantic analyses of speech in psychosis. The feature list is summarized in Table 4 (for a complete list see Table S1 in SM). Semantic similarities values equal to one (for fastText and SentenceTransformers) were excluded from calculations in order not to bias the values because of the effect of repetitions.

**Table 4:** Semantic features.

| **Static variables** |  |
| --- | --- |
| **Average Euclidean distance** | Cumulative Euclidean distance between consecutive embeddings divided by the number of distances |
| **Mean, median, max, min, std** | mean, median, maximum, minimum, standard deviation of all semantic similarity values (word window = 5) |
| **p10, p25, p75, p90** | 10th, 25th, 75th, 90th percentile of the consecutive similarity curve (word window = 5) |
| **Dynamic variables (for consecutive similarities only)** |  |
| **Autocorrelation** | first, second, third lag coefficient of autocorrelation of differences between points of consecutive similarity times series |
| **Slope sign changes** | number of times the consecutive similarity time series changes the sign of the slope (normalized by the number of opportunities of change) |
| **Crossings** | number of times the consecutive similarity curve crosses the mean value (normalized by the number of possible pairs in the list) |

*Note:* The word window for fastText and BERT semantic similarities calculation was equal to 5 (1st-5th order), while only consecutive semantic similarities were calculated for sentences. Mean, median, maximum, minimum, standard deviation, percentiles were calculated for semantic similarities from the 1st to the 5th order.

### 2.5 Principal component analysis and composite index

In order to reduce the dimensionality of 117 semantic measures, while keeping its intrinsic complexity, PCA was performed, using the ‘scikit-learn’ package in Python. We identified 54 missing values across the 4 timepoints for sentence similarity autocorrelations lag 1-3, total of 54 across four timepoints; there were no other missing values. To prepare the data to perform the PCA, these missing values were imputed separately for SSD and HC groups, using Iterative Imputer with Linear Regression model as estimator. Initial PCA was performed on a data frame with 117 features, 139 observations for T1 and a random seed equal to 8. To decide on the optimal number of components to retain for the following steps, the Kaiser Criterion (Kaiser, 1960) was applied, i.e. components with Eigenvalues higher than 1 were retained. 24 principal components were initially chosen.

To obtain a composite index as a single indicator that summarizes the features for each participant, we calculated a weighted average of the first selected principal components (Nardo et al., 2005; Chao, 2017). Specifically, for each participant, the values of the first retained principal components were calculated, and the proportion of variance explained was used as the weight for calculating the average. This was performed in two steps to ensure that different aspects of semantic behavior were fairly represented. Since we do not a priori know how various semantic features relate to distinct aspects of speech, many may be correlated and capture similar dimensions. Without addressing this, an index could overrepresent certain aspects simply because more features contribute to them. To mitigate this, we first applied PCA to balance the contributions of different features and prevent overweighting.

First, a preliminary index was calculated for the initially retained components. While PCA reduces the dimensionality of the space, the number of the original features does not change. Therefore, it is useful to discard redundant variables. For that, a Spearman correlation matrix was calculated to exclude highly correlated variables (r > 0.8). From the pairs of highly correlated features, the one with lower correlation with the previously calculated index was dropped. This step helped to reduce the number of semantic features by 49, bringing the total of 117 features down to 68. The final PCA for the first timepoint was then run with this reduced number of features. 20 principal components with eigenvalues higher than 1 were retained this time and explained 78% of the total variance. The final composite index calculation was performed based on the final number of components (n = 20) and features (n = 68). For the purpose of comparability of data across four timepoints, the indices for timepoints 2 – 4 were calculated using the projection matrix from the final PCA for the first timepoint (using the same eigenvectors and same final weights). All the composite indices were normalized by z-score and scaled from 1 to 10, for interpretability. In Figure S1 of the supplementary materials, a flow-chart summarizing the calculation of the indices as well as the steps of the PCAs can be found. For the scree plot of the Eigenvalues for each principal component, including Kaiser criterion, see Figure S2.

### 2.6 Statistical analyses

Linear mixed-effects models were used to estimate group differences (SSD vs. HC) in the transition between two timepoints, to assess changes in the index over four timepoints for the SSD group only, as well as to regress the sum of TLC scores on the index. The index was log-transformed to address skewness in the distribution both as a predictor and response variable. In the first model, we examined how the interaction between group and time influenced the index, while controlling for participant-level random effects and covariates such as age, education, and sex. In the second model, time was included as a factor with forward differences coding; the indices were regressed on time as a fixed effect, participants as random effects while the same covariates of age, education and sex were added. Pearson correlation was applied to see the correlation between the TLC scores and the index. Additionally, an ordinal logistic regression model was employed to examine whether discrete changes from one score to the next could be explained by the index. Statistical analyses and visualizations were done in RStudio, version 4.4.1, and Python, version 3.12.7.

## 3. Results

### 3.1 Group differences in the composite index

We first evaluated differences in the composite indices of all the patients (acute and stable) and controls between T1 and T3 (3 months apart) (see Figure S3). Results of the mixed-effects linear regression model, which includes only acute SSD in LPoP and HC in Remora, are shown in Table 5. There was a significant group effect (p = 0.004), while the effect of time (p = 0.806) and the interaction of group and time (p = 0.061) were non-significant. There were no significant effects of sex, education or age. The plots with the model predictions are shown in Figure 1. There was a significantly lower index score in SSD compared to HC at T1 in the acute stage, but a similar score at T3 outpatient follow-up.

**Figure 1:**
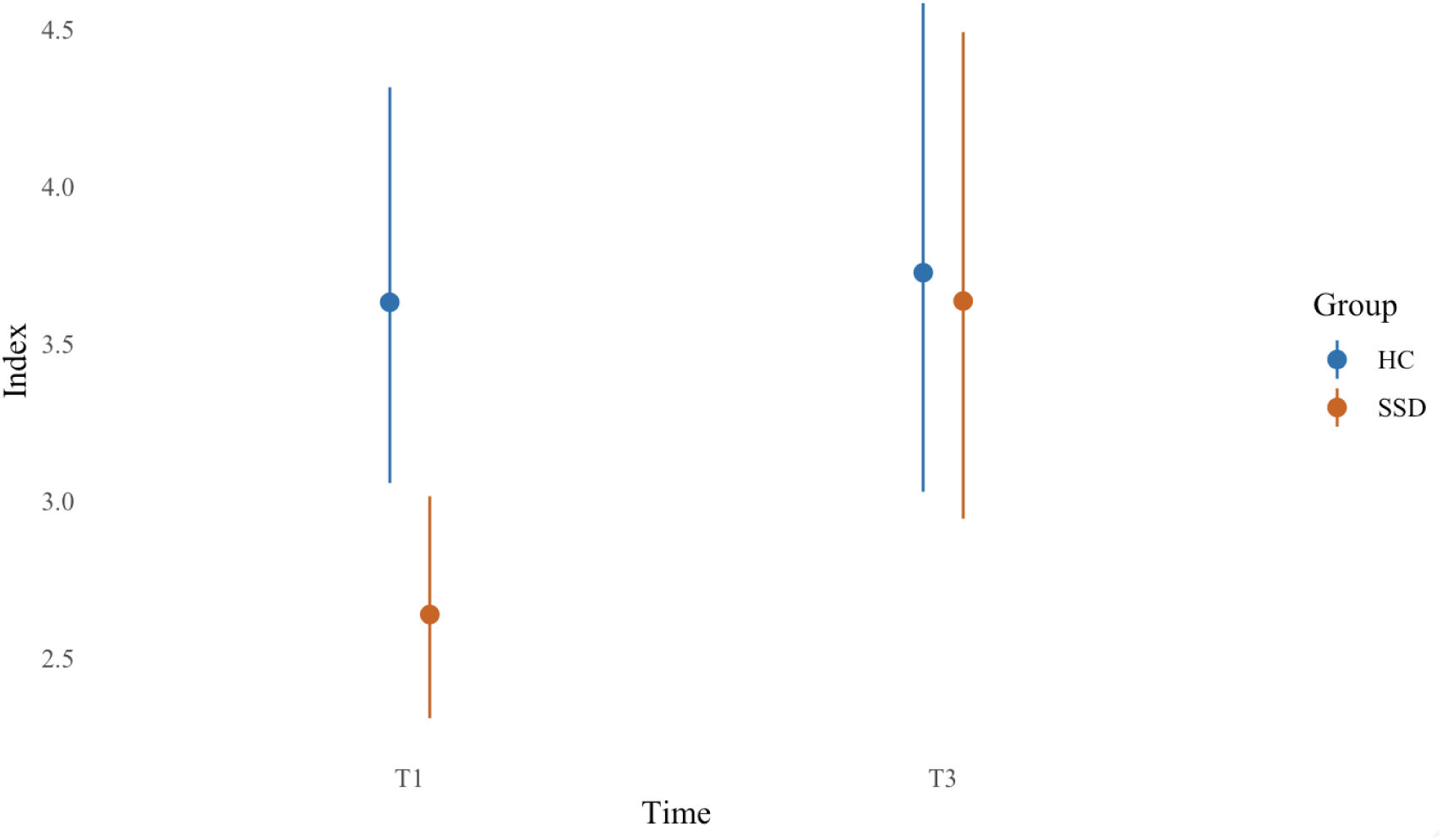
Model predictions of effect of time and group interaction on index in SSD and HC cohorts.

**Table 5:**
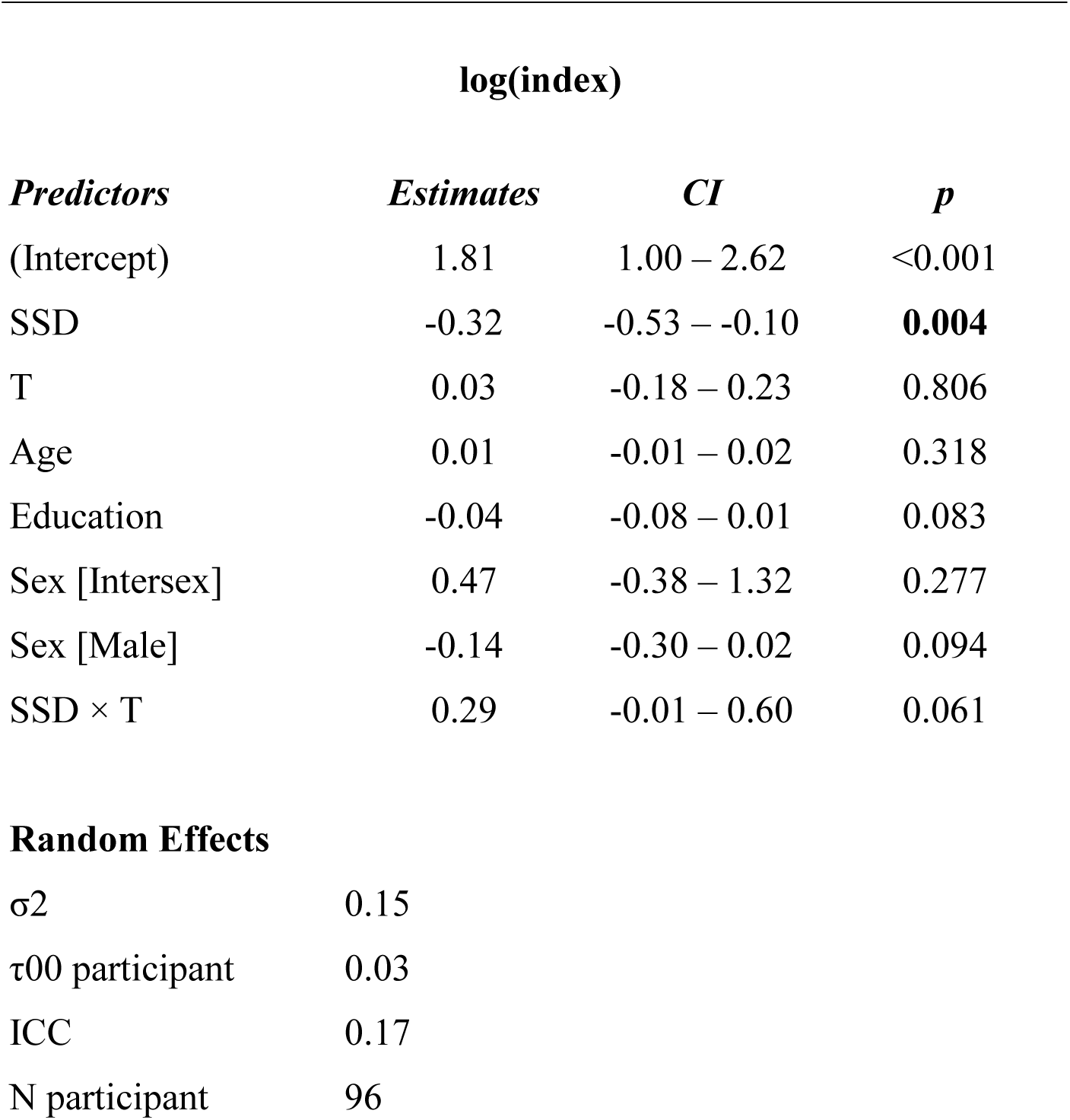
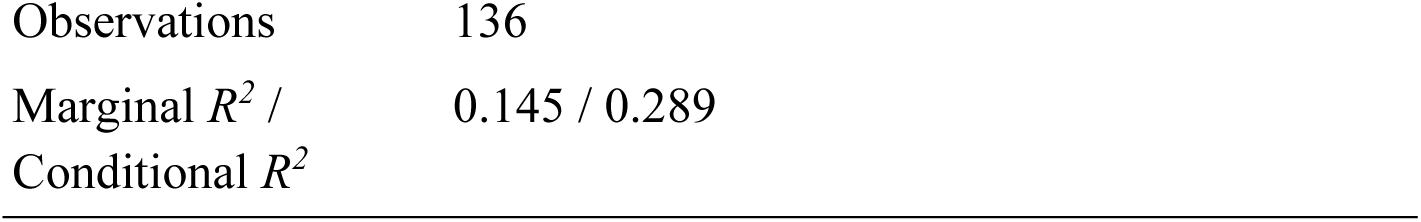
Results of the mixed-effect linear regression for LPoP SSD and Remora HCs showing the interaction of group and time on index between T1 and T3.

| **log(index)** |  |  |  |
| --- | --- | --- | --- |
| *Predictors* | *Estimates* | *CI* | *p* |
| (Intercept) | 1.81 | 1.00 – 2.62 | <0.001 |
| SSD | -0.32 | -0.53 – -0.10 | **0.004** |
| T | 0.03 | -0.18 – 0.23 | 0.806 |
| Age | 0.01 | -0.01 – 0.02 | 0.318 |
| Education | -0.04 | -0.08 – 0.01 | 0.083 |
| Sex [Intersex] | 0.47 | -0.38 – 1.32 | 0.277 |
| Sex [Male] | -0.14 | -0.30 – 0.02 | 0.094 |
| SSD × T | 0.29 | -0.01 – 0.60 | 0.061 |
| **Random Effects** |  |  |  |
| $\sigma^2$ | 0.15 | | |
| $\tau_{00}$ participant | 0.03 | | |
| ICC | 0.17 |  |  |
| N participant | 96 |  |  |

|  |  |
| --- | --- |
| Observations | 136 |
| Marginal $R^2$ /<br>Conditional $R^2$ | 0.145 / 0.289 |

### 3.2 Composite index across time

To assess the evolution of the index over a longer time frame involving four timepoints, an analysis of SSDs from LPoP-only was performed. Descriptive box plots with the changing indices across four timepoints can be found in Figure 2.

**Figure 2:**
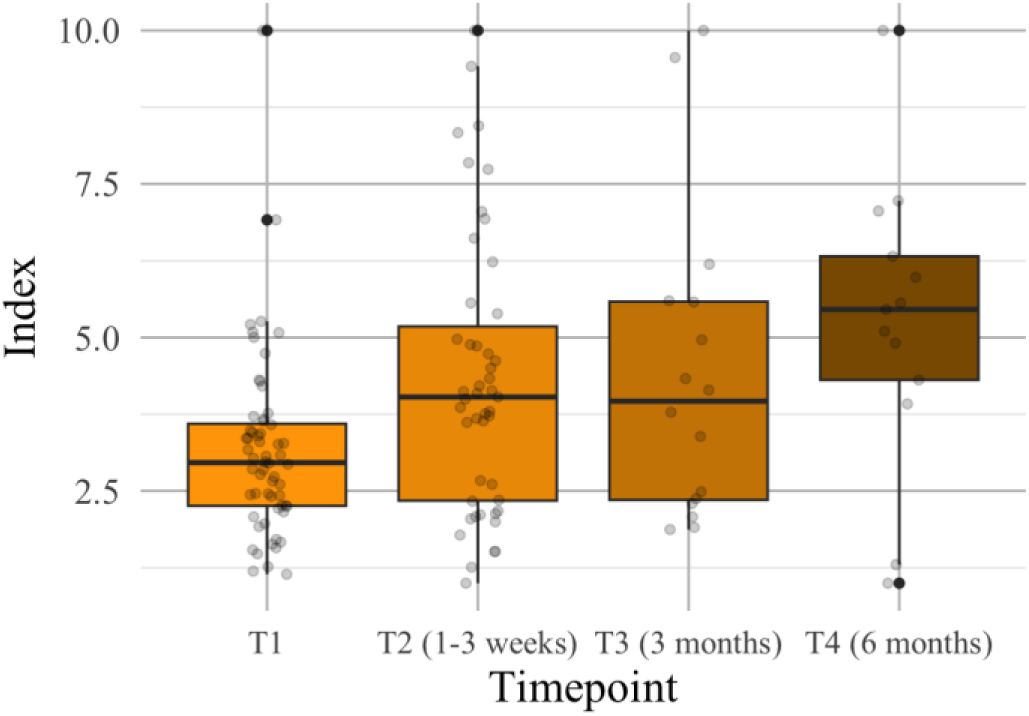
Descriptive box plots with the changing indices for LPoP SSD participants across four timepoints. Further statistical analysis of the pairwise timepoints comparisons can be found in Table S2.

The results of the mixed-effects linear model are shown in Table 6. In particular, there was a significant effect of time between T1 and T2 (p = 0.005), indicating a high increase in the index from T1 to T2. On the other hand, the time effect between T2 and T3 was non-significant (p = 0.814), indicating no significant change in the index within this period. The time effect between T3 and T4 was not significant either (p = 0.645), confirming a plateauing of the index between 3 and 6 months after the acute stage. The plot with the model predictions can be found in Figure 3.

**Figure 3:**
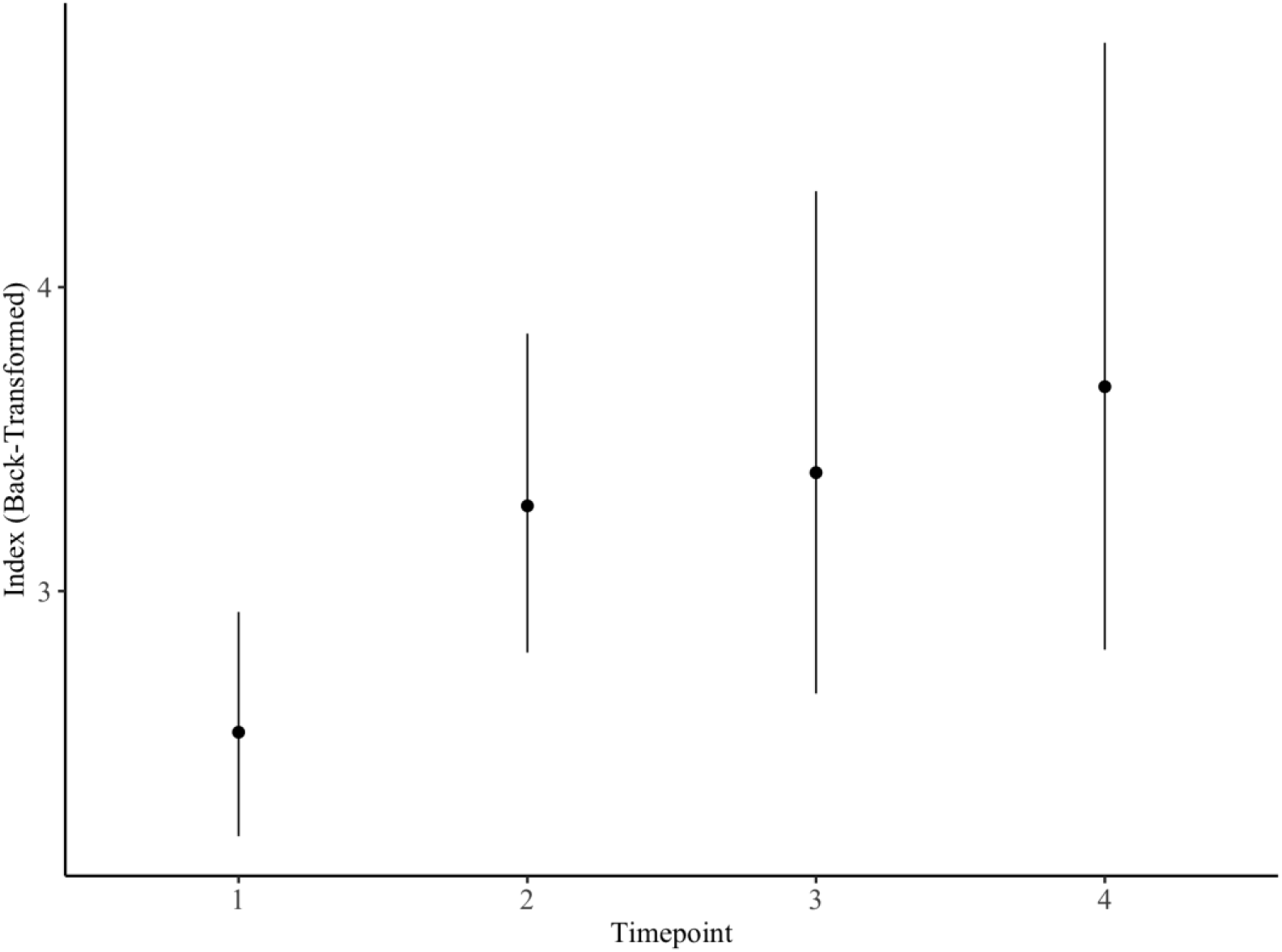
Effect of time on index in LPoP participants between T1-T2 (1-3 weeks), T2-T3 (3 months), T3-T4 (6 months).

**Table 6:**
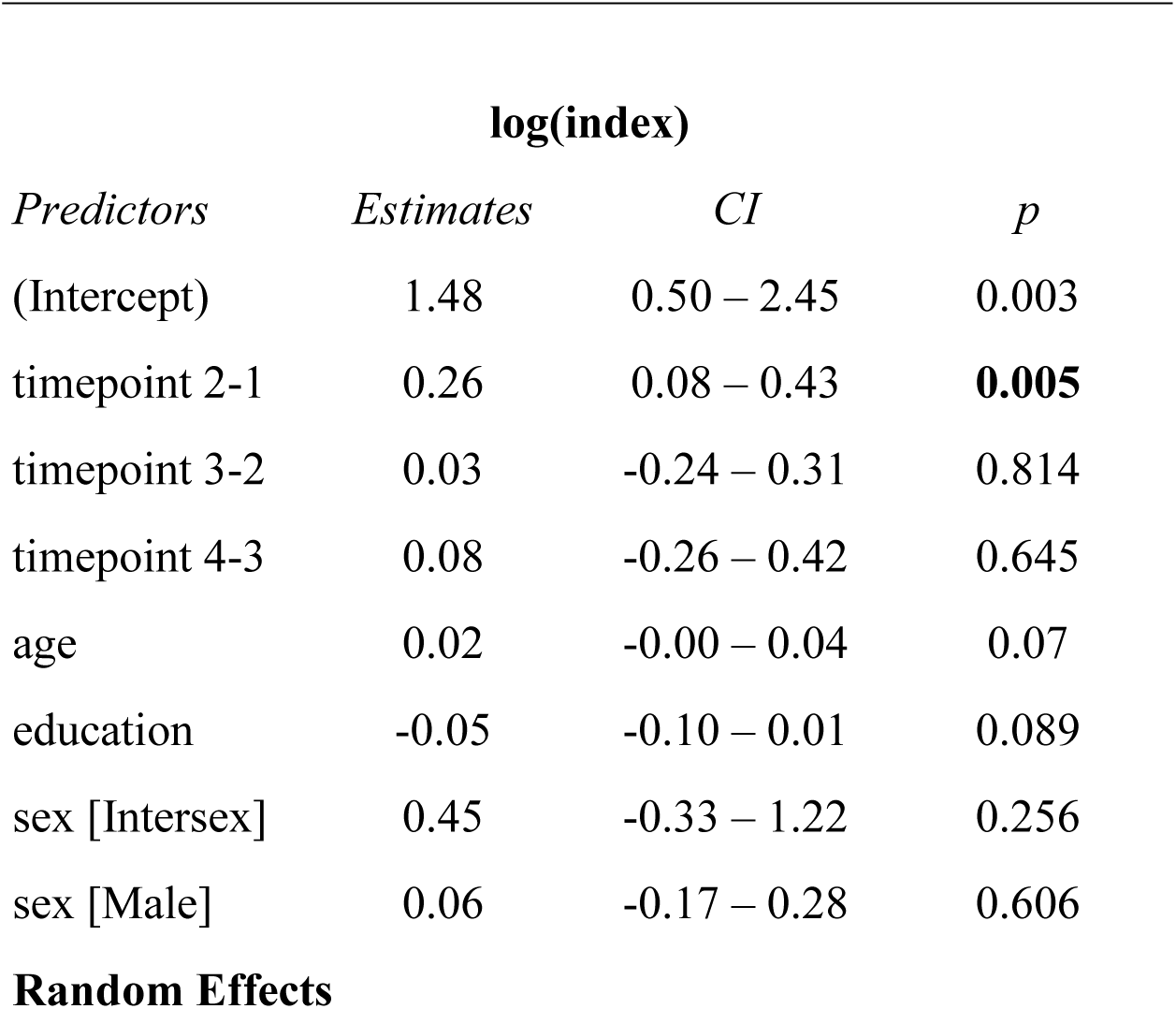
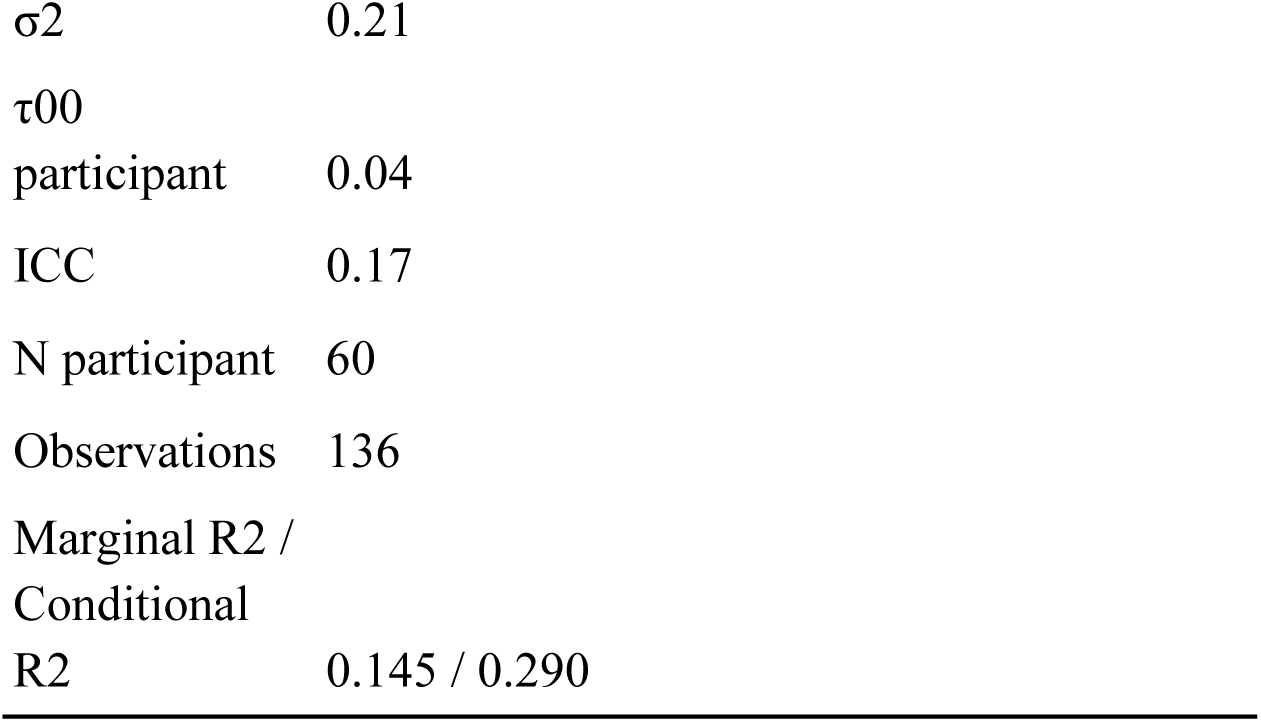
Results of the mixed-effect regression model showing the effect of time on index between the timepoints in LPoP-only cohort.

| **log(index)** |  |  |  |
| --- | --- | --- | --- |
| *Predictors* | *Estimates* | *CI* | *p* |
| (Intercept) | 1.48 | 0.50 – 2.45 | 0.003 |
| timepoint 2-1 | 0.26 | 0.08 – 0.43 | **0.005** |
| timepoint 3-2 | 0.03 | -0.24 – 0.31 | 0.814 |
| timepoint 4-3 | 0.08 | -0.26 – 0.42 | 0.645 |
| age | 0.02 | -0.00 – 0.04 | 0.07 |
| education | -0.05 | -0.10 – 0.01 | 0.089 |
| sex [Intersex] | 0.45 | -0.33 – 1.22 | 0.256 |
| sex [Male] | 0.06 | -0.17 – 0.28 | 0.606 |
| **Random Effects** |  |  |  |

|  |  |
| --- | --- |
| $\sigma^2$ | 0.21 |
| $\tau_{00}$ | |
| participant | 0.04 |
| ICC | 0.17 |
| N participant | 60 |
| Observations | 136 |
| Marginal R2 /<br>Conditional<br>R2 | 0.145 / 0.290 |

### 3.3 Relation to symptoms

The results of a mixed-effects model regressing the sum of individual TLC scores on the index across timepoints are shown in Table 7. The model explained 65.6% of the variance in the sum of TLC scores (Conditional R² = 0.656), revealing a significant effect of the index while also highlighting the variability between participants. For better visualization, Figure 4 illustrates the correlation between the index and the sum of TLC scores, with a Pearson correlation coefficient of r = –0.24 (p = 0.004). As a reference, Figure 5 shows how the TLC scores change over the four time points, considering the average value and standard deviation (A), all the transitions for individual participants (B), and only transition from T1 to T2 (C).

**Figure 4:**
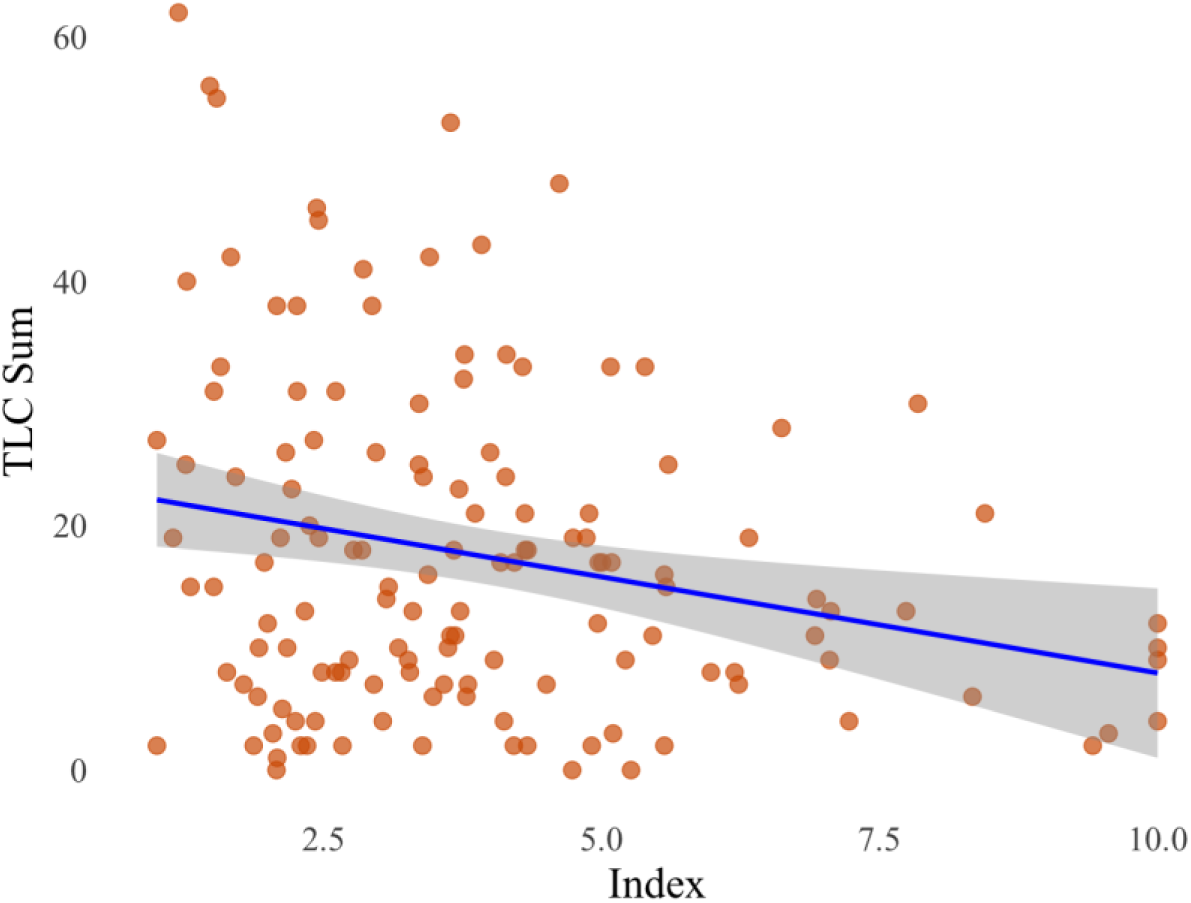
Correlation between the sum of TLC scores and the composite index.

**Figure 5:**
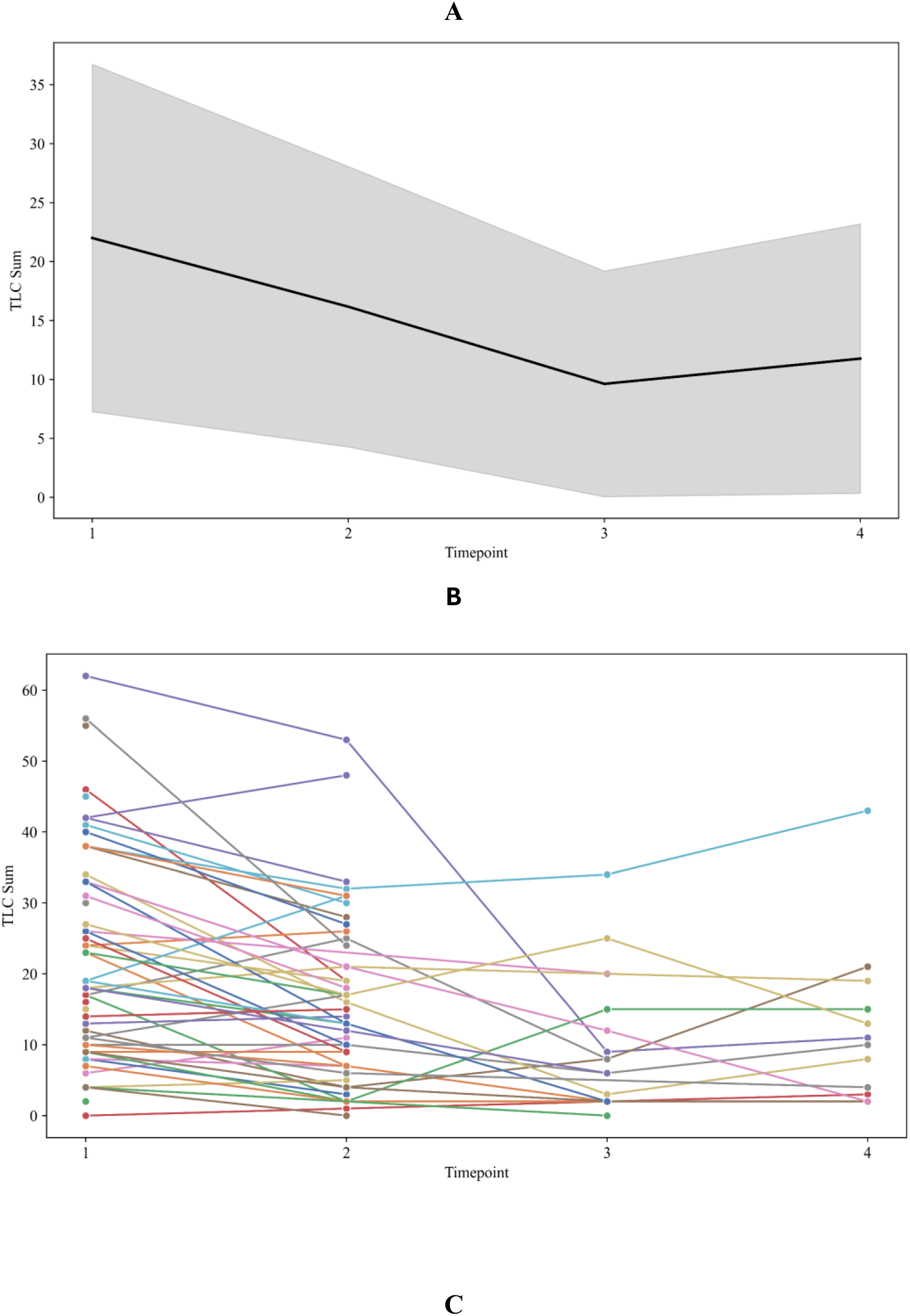
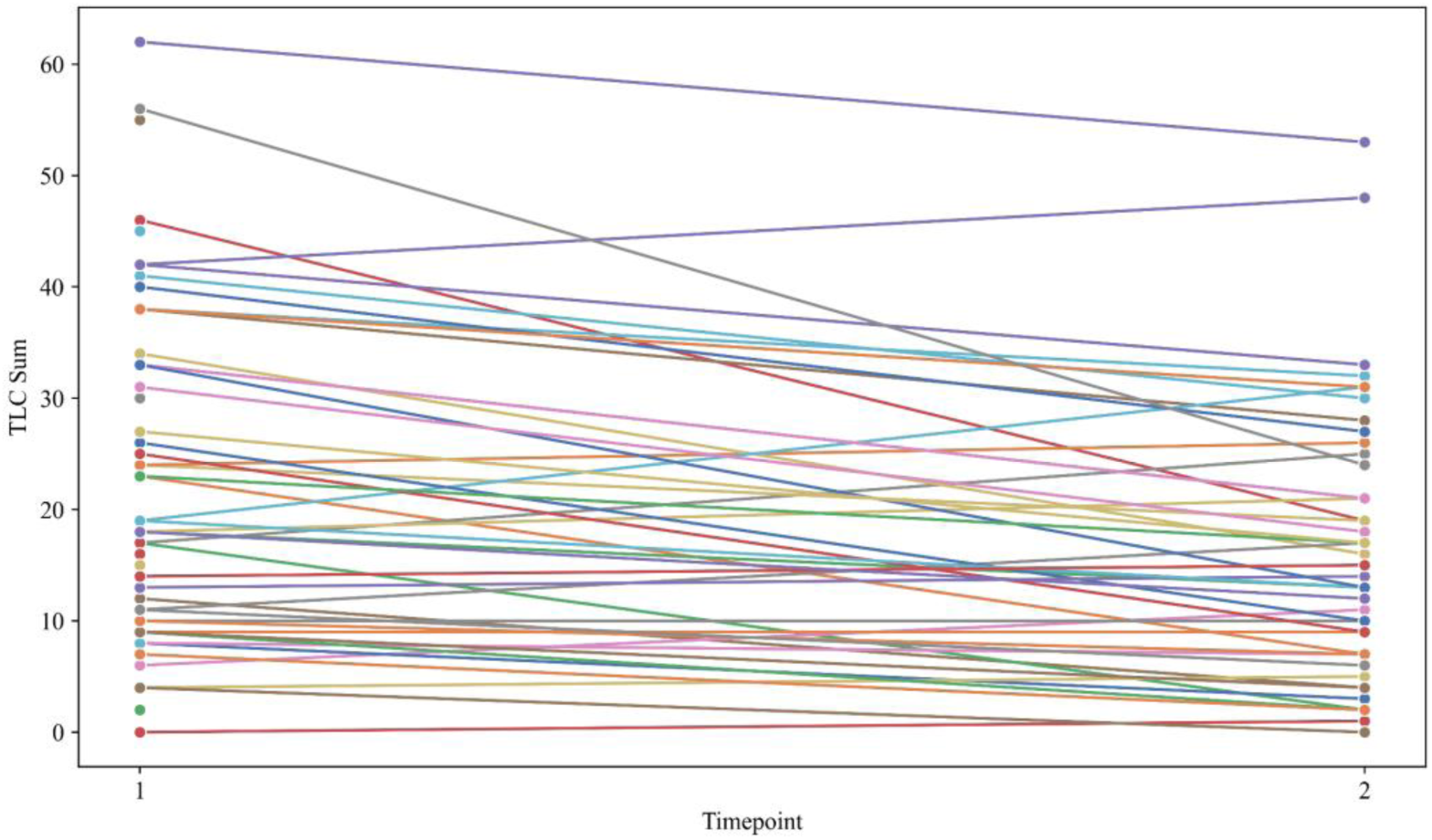
Variation in time of the sum of TLC scores.

**Table 7:** Summary of mixed linear model regression results for sum of TLC scores.

| tlc_sum |  |  |  |
| --- | --- | --- | --- |
| *Predictors* | *Estimates* | *CI* | *p* |
| (Intercept) | 39.31 | 8.93 – 69.69 | 0.012 |
| index [log] | -9.03 | -12.61 – -5.45 | **<0.001** |
| age | 0.23 | -0.38 – 0.84 | 0.457 |
| education | -1.2 | -3.02 – 0.61 | 0.192 |
| sex [Intersex] | 12.84 | -11.74 – 37.42 | 0.303 |
| sex [Male] | 0.68 | -6.37 – 7.73 | 0.849 |
| **Random Effects** |  |  |  |
| $\sigma^2$ | 70.17 | | |
| $\tau_{00}$ | 109.71 | | |
| participant |  |  |  |
| ICC | 0.61 |  |  |
| N participant | 60 |  |  |
| Observations | 136 |  |  |
| Marginal $R^2$ / | 0.118 / 0.656 | | |
| Conditional $R^2$ | | | |

To explore the relationship between the index and specific TLC subscales, we finally ran ordinal logistic regressions using the TLC loss of goal score, and the derailment score as dependent variables, as these showed the highest correlations. As seen in Tables 8-9, both regressions reveal a significant effect of the index, where a higher index is associated with a lower score. However, the sensitivity of the index differed between the subscales (referred to as ‘cuts’ below). In particular, for loss of goal, the index showed greater sensitivity to changes between scores 2 and 3. In contrast, for derailment, the effect of the index was less pronounced and mainly sensitive to changes between scores 0 and 1.

**Table 8:** Ordinal logistic regression of TLC loss of goal score.

| Variable | Coefficient | Std. Error | *z* | *p* > \|*z*\| | 95% CI |
| --- | --- | --- | --- | --- | --- |
| **log(index)** | -1.227 | 0.360 | -3.411 | 0.001 | [-1.931, -0.522] |
| **Threshold** |  |  |  |  |  |
| **Cut1 (0/1)** | -0.846 | 0.448 | -1.888 | 0.059 | [-1.723, 0.032] |
| **Cut2 (1/2)** | -0.478 | 0.237 | -2.012 | 0.044 | [-0.943, -0.012] |
| **Cut3 (2/3)** | 0.877 | 0.204 | 4.290 | 0.000 | [0.476, 1.278] |

**Table 9:**
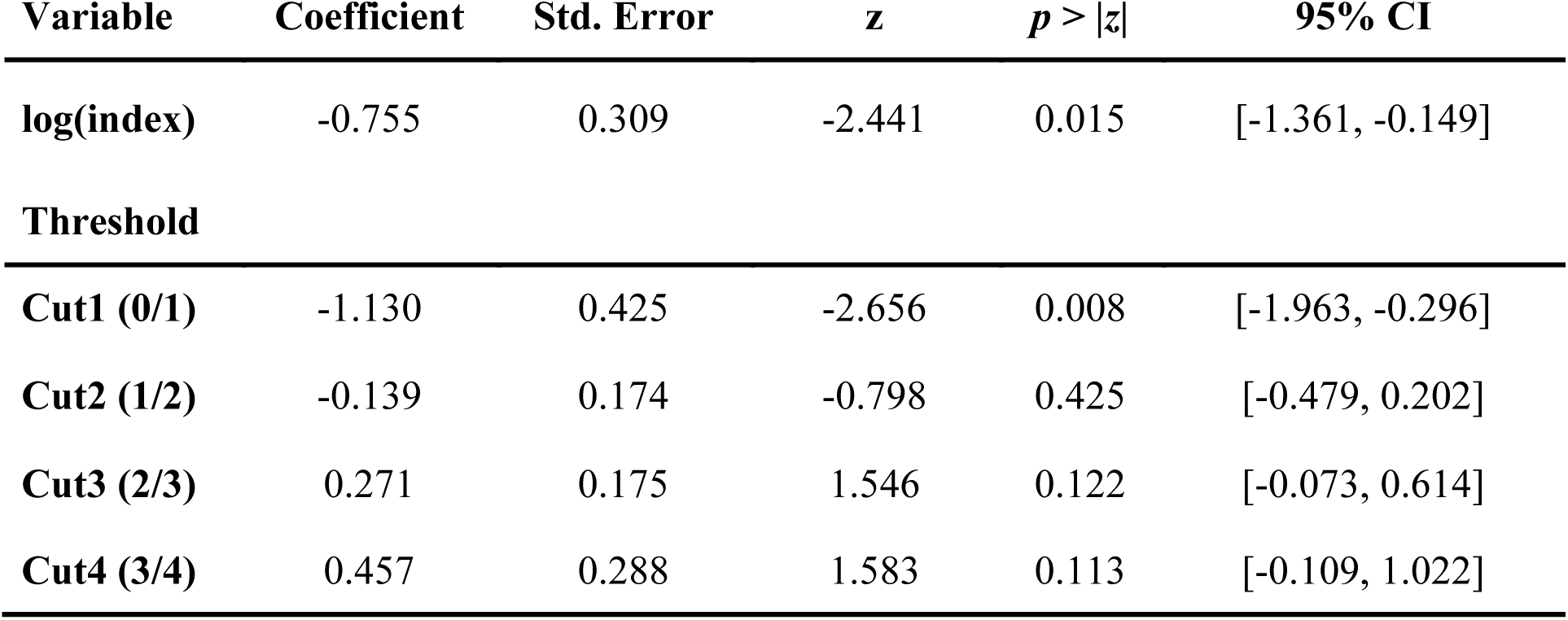
Ordinal logistic regression of TLC derailment score.

## 4. Discussion

This study aimed to define a single composite index as a summary indicator encompassing a comprehensive set of features characterizing semantic associations, intended to capture the joint behavior of these variables and their interactions across groups and time points. We have shown that this index was effective in distinguishing between groups and sensitive to changes in the symptomatic state, from acute to remission and stabilization. Additionally, it was significantly associated with the TLC, capturing 65% of the variance in the sum of TLC scores. Each in their own way, the semantic variables in this study characterize semantic associations, which are clearly affected in SSD. While the index was intended here as an agnostic tool, which as such cannot resolve inconsistencies in the literature about the direction of the association between semantic similarity and psychosis, it thus validates the intuition of thought disorder in SSD as a ‘disturbance of associations’ (Bleuler, 1911).

At the same time, when considering all variables together, our results may also point to a ceiling explanatory power of semantic variables, as suggested by the remaining variance to be explained in the sum of individual TLC scores. This raises the question of which sources of variance we may be missing. One possible explanation is the neglect of the structural organization of semantic content. In spontaneous speech, meaning is organized beyond the word level through grammar, which mediates meaning composition at different levels of the syntactic hierarchy (Hinzen & Sheehan, 2013). Such composition gives rise to different forms of meaning not found in the lexicon, such as referential meaning to a specific man as in *a man that entered the room*. This type of meaning is also the one that allows us to bind an entity identified as *a man that entered the room* to the definite noun phrase *the man* and then the pronoun *he*, so as to identify the same entity each time. Evidence of disturbances of this grammar-mediated level of referential meaning in SSD is longstanding (Rochester & Martin, 1979; Çokal et al., 2018; Palominos et al., 2023; Çokal et al., 2023). Aspects of referential meaning are often overlooked in analyses that focus only on conceptual associations, which are generally more amenable to current NLP tools. Nonetheless, it is obvious that the organization of meaning in discourse relies on both lexical and grammatical connectivity (Chomsky, 1957; Ardila, 2021). In the field of speech analysis in SSD, it is therefore crucial to integrate both levels of analysis in a theory-driven framework, which explains their relationships and predicts variability based on clinical rating scales.

The two highest correlations between the index and clinical subscales were for ‘loss of goal’ and ‘derailment’. These are symptoms observed at the discourse level, to which referential coherence is essential (Kuperberg, 2010). It is noteworthy, therefore, that the computational semantic metrics used here, which effectively capture lexical-semantic associations rather than referential meaning, still induce these correlations. This may suggest that disorders at the level of referential coherence leave a reflex at the level of lexical-semantic associations as well. Although the ordinal regressions indicated that the index partially explains the symptoms of ‘loss of goal’ and ‘derailment’, it is noteworthy that the index offers varying levels of evidence depending on the discrete transitions between scores. In particular, the analysis shows that the index is particularly sensitive to the opposite ends of these two symptom scores, indicating a more pronounced explanatory power at the extremes. We emphasize that referential coherence, while an important aspect of discourse, is only one component of overall discourse coherence. This aligns with the view that discourse-level disruptions involve a complex interplay of referential, contextual, and pragmatic factors.

The composite index also contributes to understanding how the different variables separately capture distinct aspects of semantic associations. Thus, from the top loading features of the PCA for T1 (Figure S4), their contribution to each principal component can be discerned. For example, in Figure S4, we observe the 25th percentile of consecutive semantic similarity using fastText (ft_1_p25) correlating positively with the first component, thereby increasing the value of the index. This implies that the lowest similarity values between words are likely higher in healthy controls. However, this finding does not provide information about other variables related to word similarity, which may also be correlated. In particular, there are many BERT-related variables among the top loaders too, in contrast with only two fastText variables, which emphasizes their contributions to the analysis. This could be due to 1) more sensitivity of BERT variables to capture nuances in similarities because of contextual representations, or 2) fastText variables being more correlated to each other, capturing less variance together, and a unique semantic aspect as they are static embeddings. Regardless of that, it is essential to explore explanatory criteria that illuminate these interactions and enhance the interpretability of the variables. Moreover, work needs to continue that has already shown variables from these different language models to pick up changes in brain structure and function, suggesting the potential of spontaneous speech in acting as an almost freely available readout of neurocognitive changes (Palaniyappan et al., 2019; Limongi et al., 2023; He et al., 2024b; Alonso-Sánchez et al., 2024).

### Limitations and future research

First, sample sizes diminished with time points of assessment, raising the possible doubt of selective dropping-out effects. Conceivably, the more poorly functioning participants were the ones lost to follow-up, distorting our results. Second, as described in the methods, the focus of the variables was on local semantic associations; that is, the calculations of semantic similarity were performed for consecutive or proximate words (e.g., second, third, fourth and fifth order), rather than for larger word windows or global associations across all words. Third, while we addressed the associations between both lexical-conceptual items and sentences, we must consider that different sentences may also vary in length. It is unclear to what extent this variation might influence results, particularly if there are differences in complexity and length between groups. Fourth, refinements of the index calculation should be tested: for instance, use of a different criterion to select the number of components. Fifth, more observations are needed both for initial calculation of the index and for each time point for better model predictions. We here made the methodological decision to combine two cohorts so as to maximize sample size and capture the full variability available across different groups. While this increased the clinical heterogeneity (e.g., including both inpatients and outpatients), future research is needed to determine whether results would hold when the index is calculated in an acute only sample. Finally, it is unclear whether there is a memory effect in the Remora cohort as the same picture was used in two time points, while all pictures were different for LPoP participants. Further research should explore different types of tasks, test index cross-linguistically and expand the methodology to encompass broader aspects of semantic behavior including referential meaning.

### Conclusions

Computational tools are now available that allow characterizing how speakers traverse semantic spaces, along multiple dimensions. This high dimensionality of the semantic variable space can be captured in the form of a single semantic index revealing clinically meaningful variation. This shows promise for using this index, possibly along with others capturing more aspects of variance, for practical purposes such as tracking clinically significant changes. Further research should explore different types of tasks, test the index cross-linguistically and expand the methodology to encompass broader aspects of semantic behavior including referential meaning.

## Code availability

The code used to support these findings is available from the corresponding author upon reasonable request.

## Data availability

Clinical and derived features used in these analyses are available upon reasonable request to the corresponding author. For raw transcripts from Remora please contact ST.

## Acknowledgements

We are grateful to the participants for their contributions, as well as Sarah Berretta, Leily Behbehani, William Simpson, and Jessica Robin.

## Funding

This work was supported by the European Union (GA 101080251 – TRUSTING). Views and opinions expressed are however those of the authors only and do not necessarily reflect those of the European Union or the Agency. Neither the European Union nor the granting authority can be held responsible for them. Data collection for the LPoP sample was provided by Winterlight Labs, Inc. SXT is supported by the Brain and Behavior Research Foundation Young Investigator Grant and NIH K23 MH130750.

## Conflicts of interest

S.X.T. owns equity and serves on the board and as a consultant for North Shore Therapeutics, received research funding and serves as a consultant for Winterlight Labs, is on the advisory board and owns equity for Psyrin, and serves as a consultant for Catholic Charities Neighborhood Services and LB Pharmaceuticals. M.S. is a full-time employee of Cambridge Cognition.

## Author contributions

C.P.: Conceptualization, Investigation, Data Analysis, Methodology, Writing – Original Draft, Writing – Review & Editing, Formal analysis, Project administration. M.K.: Investigation, Data Analysis, Formal analysis, Methodology, Writing – Original Draft, Writing – Review & Editing. A.N.: Resources, Data Curation, Writing – Review & Editing. M.S.: Writing – Review & Editing. P.H.: Writing – Review & Editing, Funding Acquisition. I.S.: Writing – Review & Editing, Funding Acquisition. S.T.: Resources, Data Curation, Writing – Review & Editing. W.H.: Conceptualization, Writing – original draft, Writing – Review & Editing, Funding acquisition, Supervision.

## Notes

### Competing Interest Statement

Sunny Tang reports financial support was provided by North Shore Therapeutics, Winterlight Labs, Psyrin, Catholic Charities Neighborhood Services, LB Pharmaceuticals. Michael Spilka reports a relationship with Cambridge Cognition Ltd that includes: employment. Other authors declare that they have no known competing financial interests or personal relationships that could have appeared to influence the work reported in this paper.

### Author Declarations

The IRB of the Feinstein Institutes for Medical Research gave ethical approval for this work (#20-0128, #20-0289, & #20-0460)

## References

1. Abdi, H., & Williams, L. J. (2010). Principal component analysis. In Wiley Interdisciplinary Reviews: Computational Statistics (Vol. 2, Issue 4, pp. 433–459). 10.1002/wics.101.

2. Alonso-Sánchez, M. F., Limongi, R., Gati, J., & Palaniyappan, L. (2023). Language network self-inhibition and semantic similarity in first-episode schizophrenia: A computational-linguistic and effective connectivity approach. Schizophrenia research, 259, 97–103. 10.1016/j.schres.2022.04.007.

3. Alonso-Sánchez, M. F., Hinzen, W., He, R., Gati, J., & Palaniyappan, L. (2024). Perplexity of utterances in untreated first-episode psychosis: an ultra–high field MRI dynamic causal modelling study of the semantic network. Journal of Psychiatry and Neuroscience, 49(4), E252–E262.

4. Andreasen, N. C. (1979). Thought, Language and Communication Disorders: I. Clinical Assessment, Definition of Terms and Evaluation of Their Reliability. Archives of General Psychiatry, 36, 1315–1321 10.1001/archpsyc.1979.01780120045006.

5. Andreasen, N. C. (1986). Scale for the Assessment of Thought, Language, and Communication (TLC) (Vol. 12, Issue 3). https://academic.oup.com/schizophreniabulletin/article/12/3/473/1866041.

6. Ardila, A. (2021). Grammar in the brain: Two grammar subsystems and two agrammatic types of aphasia. Journal of Neurolinguistics, 58, 100960.

7. Arslan, B., Kizilay, E., Verim, B., Demirlek, C., Dokuyan, Y., Turan, Y. E., … & Bora, E. (2024). Automated linguistic analysis in speech samples of Turkish-speaking patients with schizophrenia-spectrum disorders. Schizophrenia Research, 267, 65–71.

8. Bedi, G., Carrillo, F., Cecchi, G. A., Slezak, D. F., Sigman, M., Mota, N. B., Ribeiro, S., Javitt, D. C., Copelli, M., & Corcoran, C. M. (2015). Automated analysis of free speech predicts psychosis onset in high-risk youths. Npj Schizophrenia, 1(1), 15030. 10.1038/npjschz.2015.30.

9. Bleuler E. (1911). Dementia Praecox oder Gruppe der Schizophrenien. Leipzig, Germany: Deuticke.

10. Bojanowski, P., Grave, E., Joulin, A., & Mikolov, T. (2017). Enriching word vectors with subword information. Transactions of the association for computational linguistics, 5, 135–146. 10.1162/tacl_a_00051.

11. Çabuk, T., Sevim, N., Mutlu, E., Yağcıoğlu, A. E. A., Koç, A., & Toulopoulou, T. (2024). Natural language processing for defining linguistic features in schizophrenia: A sample from Turkish speakers. Schizophrenia Research, 266, 183–189. 10.1016/j.schres.2024.02.026.

12. Chao, Y. S., & Wu, C. J. (2017). Principal component-based weighted indices and a framework to evaluate indices: Results from the Medical Expenditure Panel Survey 1996 to 2011. PLoS ONE, 12(9). 10.1371/journal.pone.0183997.

13. Choe, E., Ha, M., Choi, S., Park, S., Jang, M., Kim, M., & Kwon, J. S. (2023). Beyond verbal fluency in the verbal fluency task: semantic clustering as a predictor of remission in individuals at clinical high risk for psychosis. Journal of Psychiatry and Neuroscience, 48(6), E414–E420. 10.1503/jpn.230074.

14. Chomsky, N. (1957). Syntactic structures. Mouton.

15. Çokal, D., Sevilla, G., Jones, W. S., Zimmerer, V., Deamer, F., Douglas, M., Spencer, H., Turkington, D., Ferrier, N., Varley, R., Watson, S., & Hinzen, W. (2018). The language profile of formal thought disorder. NPJ schizophrenia, 4(1), 18. 10.1038/s41537-018-0061-9.

16. Çokal, D., Palominos-Flores, C., Yalınçetin, B., Türe-Abacı, Ö., Bora, E., & Hinzen, W. (2023). Referential noun phrases distribute differently in Turkish speakers with schizophrenia. Schizophrenia Research, 259, 104–110.

17. Devlin, J., Chang, M.-W., Lee, K., & Toutanova, K. (n.d.). BERT: Pre-training of Deep Bidirectional Transformers for Language Understanding.

18. Elvevåg, B., Foltz, P. W., Weinberger, D. R., & Goldberg, T. E. (2007). Quantifying incoherence in speech: an automated methodology and novel application to schizophrenia. Schizophrenia research, 93(1-3), 304–316. 10.1016/j.schres.2007.03.001.

19. Figueroa-Barra, A., Del Aguila, D., Cerda, M., Gaspar, P. A., Terissi, L. D., Durán, M., & Valderrama, C. (2022). Automatic language analysis identifies and predicts schizophrenia in first-episode of psychosis. *Schizophrenia (Heidelberg*, Germany), 8(1), 53. 10.1038/s41537-022-00259-3.

20. First, M. B., & Gibbon, M. (2004). The Structured Clinical Interview for DSM-IV Axis I Disorders (SCID-I) and the Structured Clinical Interview for DSM-IV Axis II Disorders (SCID-II). In M. J. Hilsenroth & D. L. Segal (Eds.), Comprehensive handbook of psychological assessment, Vol. 2. Personality assessment (pp. 134–143). John Wiley & Sons, Inc.

21. Firth, J.R. (1957). Applications of General Linguistics. Transactions of the Philological Society, 56, 1–14.

22. Grave, E., Bojanowski, P., Gupta, P., Joulin, A., & Mikolov, T. (2018). Learning Word Vectors for 157 Languages (arXiv:1802.06893). arXiv. http://arxiv.org/abs/1802.06893.

23. Harris Z.S. (1954). Distributional Structure, WORD, 10:2–3, 146-162, DOI: 10.1080/00437956.1954.11659520.

24. He, R., Palominos, C., Zhang, H., Alonso-Sánchez, M. F., Palaniyappan, L., & Hinzen, W. (2024a). Navigating the semantic space: Unraveling the structure of meaning in psychosis using different computational language models. Psychiatry Research, 333, 115752.

25. He, R., Ortiz-García de la Foz, V., Fernández Cacho, L. M., Homan, P., Sommer, I., Ayesa-Arriola, R., & Hinzen, W. (2024b). Task-voting for schizophrenia spectrum disorders prediction using machine learning across linguistic feature domains. medRxiv, 2024-08.

26. Hinzen, W. & Sheehan, M. (2013). The Philosophy of Universal Grammar. Oxford University Press. 10.1093/acprof:oso/9780199654833.001.0001.

27. Jolliffe, I. T. (2002). Mathematical and statistical properties of sample principal components. In: Principal Component Analysis. Springer Series in Statistics. Springer, New York. 10.1007/0-387-22440-8_3.

28. Kaiser, H. F. (1960). The application of electronic computers to factor analysis. Educational and Psychological Measurement, 20, 141–151. 10.1177/001316446002000116.

29. Kuperberg, G. R. (2010). Language in schizophrenia part 1: An introduction. Language and Linguistics Compass, 4 (8), 576–589.

30. Liang, L., Silva, A.M., Jeon, P., Ford, S.D., MacKinley, M., Théberge, J., Palaniyappan, L. (2022). Widespread cortical thinning, excessive glutamate and impaired linguistic functioning in schizophrenia: A cluster analytic approach. Front. Hum. Neurosci. 16:954898. doi: 10.3389/fnhum.2022.954898.

31. Limongi, R., Silva, A. M., Mackinley, M., Ford, S. D., & Palaniyappan, L. (2023). Active inference, epistemic value, and uncertainty in conceptual disorganization in first-episode schizophrenia. Schizophrenia Bulletin, 49(Supplement_2), S115–S124. 10.1093/schbul/sbac125.

32. Matharaarachchi, S., Domaratzki, M., & Muthukumarana, S. (2021). Assessing feature selection method performance with class imbalance data. Machine Learning with Applications, 6, 100170. 10.1016/j.mlwa.2021.100170.

33. Mikolov, T., Sutskever, I., Chen, K., Corrado, G. S., & Dean, J. (2013a). Distributed representations of words and phrases and their compositionality. Advances in neural information processing systems, 26.

34. Mikolov, T., Corrado, G., Chen, K. and Dean, J. (2013b). Efficient Estimation of Word Representations in Vector Space. Proceedings of the Workshop at ICLR, Scottsdale, 2-4 May 2013, 1-12.

35. Miller, G. A., & Charles, W. G. (1991). Contextual correlates of semantic similarity. Language and Cognitive Processes, 6(1), 1–28. 10.1080/01690969108406936.

36. Morgan, E., Arnold, M., Camargo, M. C., Gini, A., Kunzmann, A. T., Matsuda, T., Meheus, F., Verhoeven, R. H. A., Vignat, J., Laversanne, M., Ferlay, J., & Soerjomataram, I. (2022). The current and future incidence and mortality of gastric cancer in 185 countries, 2020-40: A population-based modelling study. EClinicalMedicine, 47, 101404. 10.1016/j.eclinm.2022.101404.

37. Nardo, M., Saisana, M., Saltelli, A., Tarantola, S., Hoffman, A., & Giovannini, E. (2005). OECD Statistics Working Papers 2005/03 Handbook on Constructing Composite Indicators: Methodology and User Guide. 10.1787/533411815016.

38. Palaniyappan, L., Mota, N.B., Oowise, S., Balain, V., Copelli, M., Ribeiro, S., Liddle, P.F. (2019). Speech structure links the neural and socio-behavioural correlates of psychotic disorders. Prog Neuropsychopharmacol Biol Psychiatry. Jan 10;88:112-120. doi: 10.1016/j.pnpbp.2018.07.007. Epub 2018 Jul 11. PMID: 30017778.

39. Palaniyappan, L. (2022). Dissecting the neurobiology of linguistic disorganisation and impoverishment in schizophrenia. In Seminars in cell & developmental biology (Vol. 129, pp. 47-60). Academic Press.

40. Palominos, C., Figueroa-Barra, A., & Hinzen, W. (2023). Coreference delays in psychotic discourse: widening the temporal window. Schizophrenia Bulletin, 49(Supplement_2), S153–S162.

41. Palominos, C., He, R., Frölich, K., Mülfarth, R., Seuffert, S., Sommer, I. E., Homan, P., Kircher, T., Stein, F., Hinzen, W. (2024). Approximating the semantic space: word embedding techniques in psychiatric speech analysis. Schizophrenia, in press.

42. Parola, A., Lin, J. M., Simonsen, A., Bliksted, V., Zhou, Y., Wang, H., Inoue, L., Koelkebeck, K., & Fusaroli, R. (2023). Speech disturbances in schizophrenia: Assessing cross-linguistic generalizability of NLP automated measures of coherence. Schizophrenia Research, 259, 59–70. 10.1016/j.schres.2022.07.002.

43. Pennington, J., Socher, R., & Manning, C. D. (2014, October). Glove: Global vectors for word representation. In Proceedings of the 2014 conference on empirical methods in natural language processing (EMNLP) (pp. 1532–1543).

44. Pintos, A. S., Hui, C. L. M., de Deyne, S., Cheung, C., Ko, W. T., Nam, S. Y., Chan, S. K. W., Chang, W. C., Lee, E. H. M., Lo, A. W. F., Lo, T. L., Elvevåg, B., & Chen, E. Y. H. (2022). A Longitudinal Study of Semantic Networks in Schizophrenia and other Psychotic Disorders Using the Word Association Task. Schizophrenia Bulletin Open, 3(1). 10.1093/schizbullopen/sgac054.

45. Rochester, S., & Martin, J. R. (1979). Crazy talk: A study of the discourse of schizophrenic speakers. New York: Plenum. 229 pages.

46. Reimers, N., & Gurevych, I. (2019). Sentence-bert: Sentence embeddings using siamese bert-networks. arXiv preprint arXiv:1908.10084.

47. Roelofs, A. (2018). A unified computational account of cumulative semantic, semantic blocking, and semantic distractor effects in picture naming. Cognition, 172, 59–72.

48. Sharpe V., Mackinley M., Eddine S.N., Wang L., Palaniyappan L., Kuperberg G.R. (2024). GPT-3 reveals selective insensitivity to global vs. local linguistic context in speech produced by treatment-naïve patients with positive thought disorder. bioRxiv, 2024-07. doi: 10.1101/2024.07.08.602512

49. Tang, S. X., Kriz, R., Cho, S., Park, S. J., Harowitz, J., Gur, R. E., Bhati, M. T., Wolf, D. H., Sedoc, J., & Liberman, M. Y. (2021). Natural language processing methods are sensitive to sub-clinical linguistic differences in schizophrenia spectrum disorders. Npj Schizophrenia, 7(1). 10.1038/s41537-021-00154-3.

50. Tang, S., Hänsel, K., Cong, Y., Berretta, S., Cho, S., Nikzad, A., … & Liberman, M. (2022). Dimensions of Speech and Language Disturbance in Psychosis and Computational Linguistic Markers. Biological Psychiatry, 91(9), S49–S50.

51. Tang, S. X., Cong, Y., Nikzad, A. H., Mehta, A., Cho, S., Hänsel, K., … & Malhotra, A. K. (2023). Clinical and computational speech measures are associated with social cognition in schizophrenia spectrum disorders. Schizophrenia Research, 259, 28–37.

52. Voppel, A., De Boer, J., Brederoo, S., Schnack, H., & Sommer, I. (2021). Quantified language connectedness in schizophrenia-spectrum disorders. Psychiatry Research, 304, 114130. 10.1016/j.psychres.2021.114130.

