## Supplementary Material for "A single composite index of semantic behavior tracks symptoms of psychosis over time"

**Supplementary Materials**

**Customized list of stop words in spaCy.**

Added to spaCy's default stop words:

*nlp.Defaults.stop_words |= {'alright', 'hm', 'kinda', 'lo', 'maybe', 'm', 'mhm', 'mm', 'mmm', "n't", 'oh', 'ok', 'okay', 'okey', 'uh', 'um', 've', 'w', 'yea', 'yeah', 'yep', 'yes'}*

Removed from spaCy's default stop words:

*nlp.Defaults.stop_words -= {'already', 'always', 'amount', 'bottom', 'call', 'eleven', 'empty', 'fifty', 'first', 'five', 'four', 'get', 'give', 'go', 'hundred', 'keep', 'last', 'made', 'make', 'many', 'move', 'name', 'never', 'next', 'nine', 'one', 'others', 'part', 'serious', 'several', 'show', 'six', 'sixty', 'sometimes', 'take', 'ten', 'third', 'three', 'together', 'top', 'twelve', 'twenty', 'two', 'various', 'whole'}*

A list of the semantic features is represented in Table S1. These features aim to assess semantic similarity between words or sentences using three different models, fastText and BERT at word level, and SentenceTransformers at sentence level. Statistical measures such as the mean, median, maximum (max), minimum (min), standard deviation (std), and percentiles aim to provide insights into the distribution of semantic similarity values across different orders. Dynamic measures such as autocorrelation coefficients (autocor_lag1-3), slope sign changes ratio (ssc_ratio), and crossings ratio (crossings_ratio) aim to capture the temporal structure and variability of semantic similarity patterns over a given speech.

**Table S1:** List of semantic features.

| **Semantic Features** | |
|---|---|
| {model name}_semsim | Cosine semantic similarity between words or sentences. Model names: 'ft' for fastText, 'bert' for BERT, 'sbert' for SentenceTransformers. |
| semsim_1 | 1st order semantic similarity between two consecutive words/sentences |
| semsim_2 | 2nd order semantic similarity between words separated by one other word |
| semsim_3 | 3rd order semantic similarity between words separated by two other words |

| | |
|---|---|
| semsim_4 | 4th order semantic similarity between words separated by three other words |
| semsim_5 | 5th order semantic similarity between words separated by four other words |
| mean, median, max, min, std | Mean, median, maximum, minimum, standard deviation of semantic similarities of different orders |
| p10, p25, p75, p90 | 10th, 25th, 75th, 90th percentile of the consecutive similarity curve calculated for semantic similarities of different orders |
| autocor_lag{1-3} | First, second, third lag coefficient of autocorrelation of differences between points of consecutive similarity times series |
| ssc_ratio | Slope sign changes ratio: the number of times the consecutive similarity time series changes the sign of the slope (normalized by the number of opportunities of change) |
| crossings_ratio | Crossings ratio: the number of times the consecutive similarity curve crosses the mean value (normalized by the number of possible pairs in the list) |

**Figure S1:** Flow-chart overview for index calculation and PCA steps.

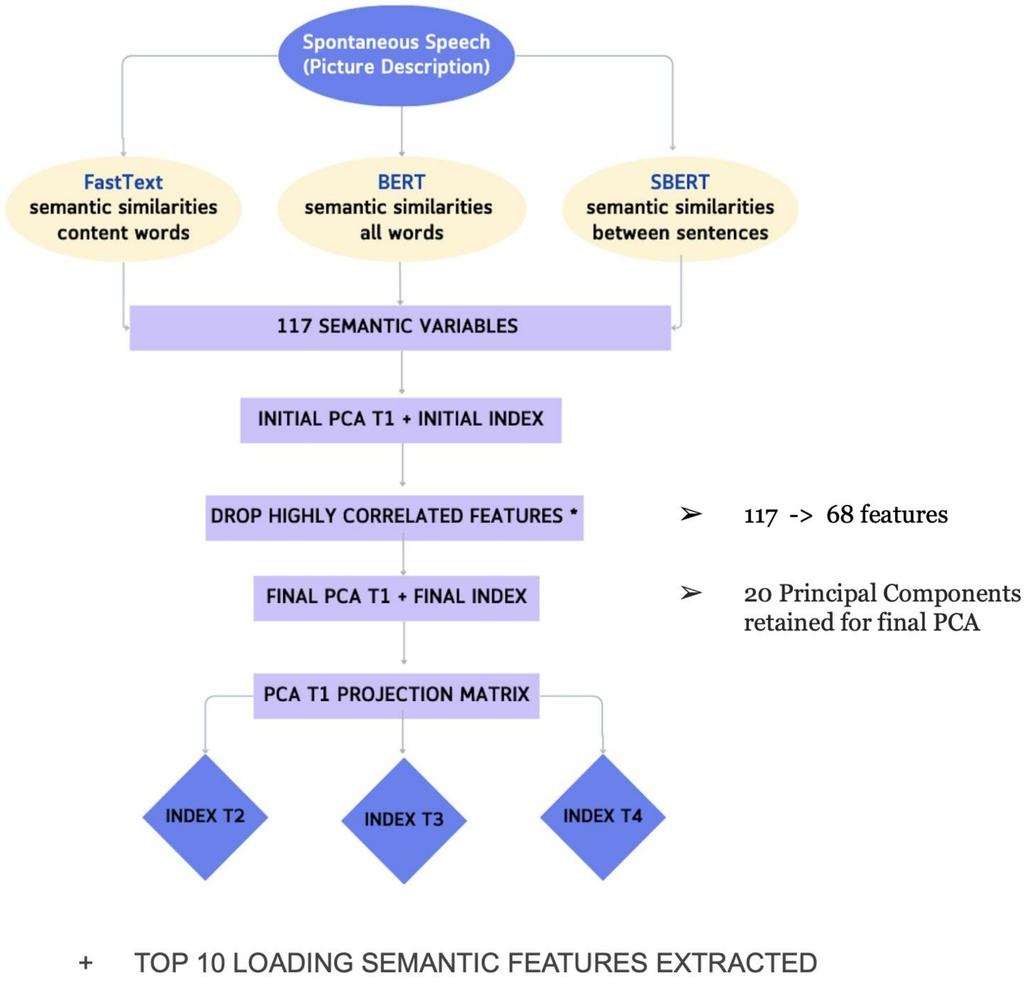


**Figure S2:** Scree plot of Eigenvalues for each Principal Component, including Kaiser criterion.

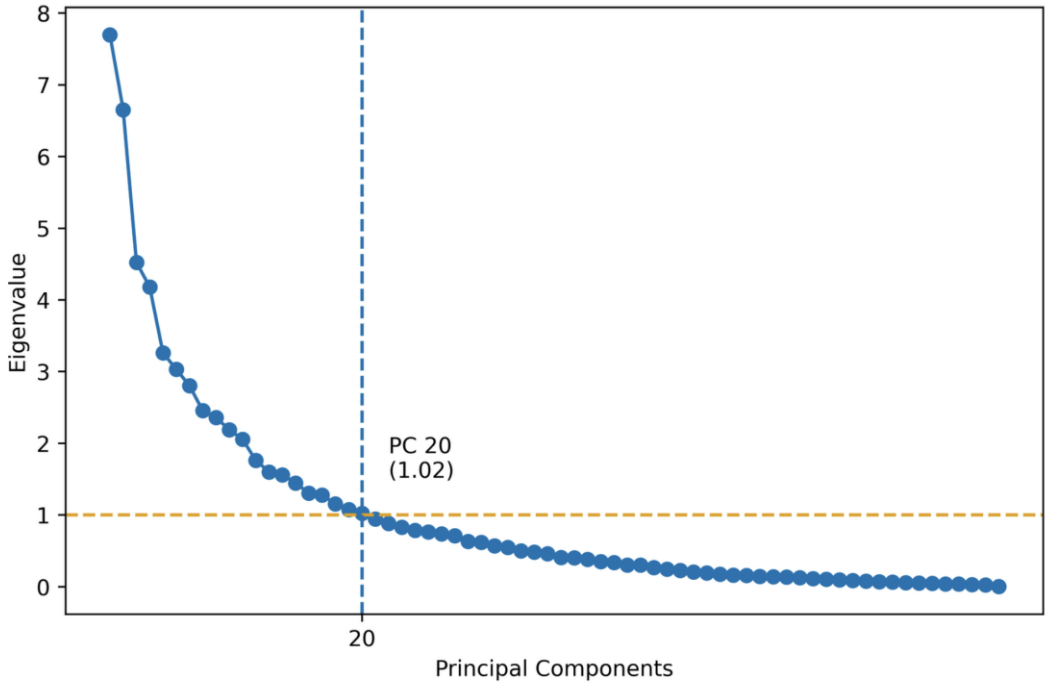


**Figure S3**: Differences in index between T1, which included a mix of acute and more stable patients of LPoP and Remora, and T3 with all patients at a stable stage. Mann-Whitney U Tests showed a statistically significant difference between the indices of SSD and HC participants at T1 (U = 1356, p = 0.017), which disappeared at T3 (U = 393, p = 0.968).

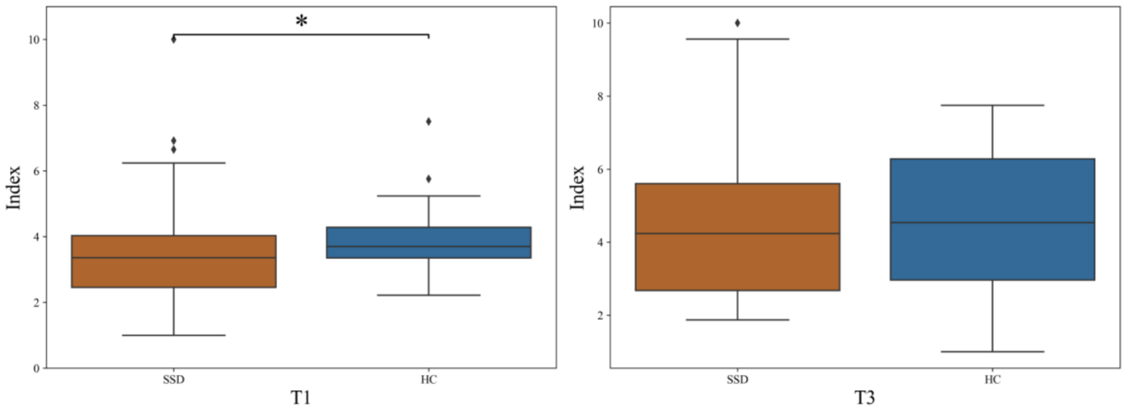


### Top loadings

The heat map of the top ten loading semantic features can be found below.

**Figure S4**: Top 10 loading features used in the PCA for T1, based on 68 semantic features and 20 principal components. Numbers from 1 to 5 correspond to the 1st - 5th order semantic similarities calculated in fastText (ft), BERT (bert) and SentenceTransformers (sbert). The top ten loading features were extracted across the final 20 components. First, the loadings were calculated by taking the transpose of the eigenvectors and multiplying it by the square root of the eigenvalues. After that they were ranked from highest to lowest based on the absolute sum of loadings for each feature and the top ten features were extracted.

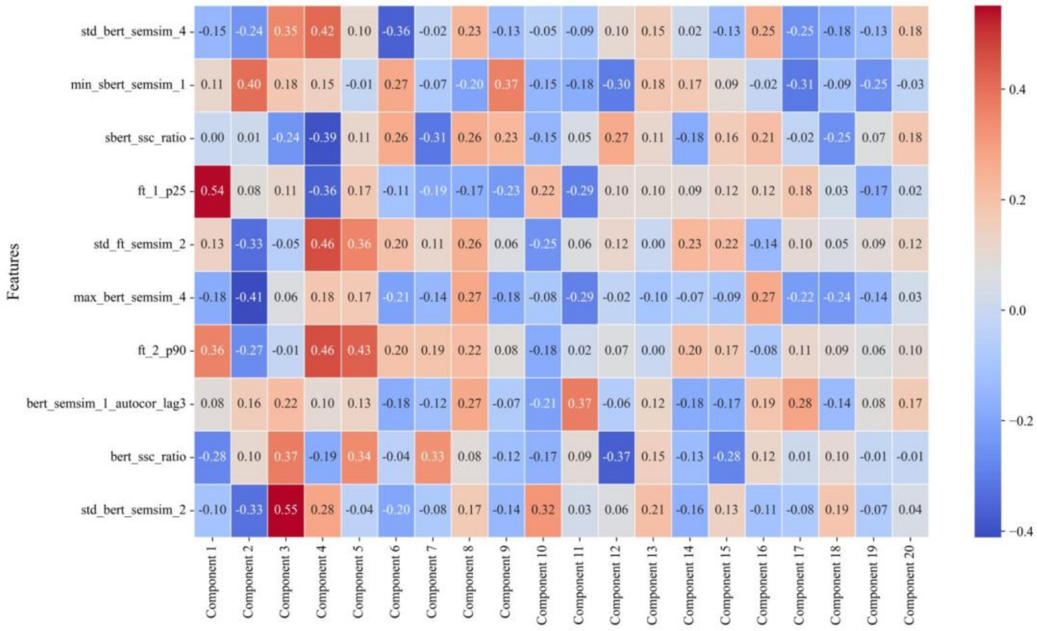


**Table S2. The pairwise comparison of differences in index for LPoP SSD participants across four timepoints (addition to Figure 3).**

An ANOVA test showed a highly significant effect of time on the index values ($F = 5.842$; $p < 0.001$). Subsequent pairwise Mann-Whitney U tests revealed significant differences between T1 vs. T2 and T1 vs. T4. No significant differences were found between other pairs of timepoints.

| **Comparison** | **U-statistic** | **p-value** |
|---|---:|---:|
| Timepoint 1 vs. 2 | 950.5 | **0.004** |
| Timepoint 1 vs. 3 | 339.5 | 0.075 |
| Timepoint 1 vs. 4 | 158.5 | **<0.001** |
| Timepoint 2 vs. 3 | 370.5 | 0.937 |
| Timepoint 2 vs. 4 | 213 | 0.099 |
| Timepoint 3 vs. 4 | 77.5 | 0.254 |
